# Automated analysis of written language in the three variants of primary progressive aphasia

**DOI:** 10.1101/2022.07.24.22277977

**Authors:** Sylvia Josephy-Hernandez, Neguine Rezaii, Amelia Jones, Emmaleigh Loyer, Daisy Hochberg, Megan Quimby, Bonnie Wong, Bradford C. Dickerson

## Abstract

Despite the important role of written language in everyday life, abnormalities in functional written communication have been sparsely investigated in Primary Progressive Aphasia (PPA). Prior studies have analyzed written language separately in the three variants of PPA – nonfluent (nfvPPA), logopenic (lvPPA), and semantic (svPPA) – but have rarely compared them to each other or to spoken language. Manual analysis of written language can be a time-consuming process. We developed a program which uses a language parser and quantifies content units (CU) and total units (U) in written language samples. The program was used to analyze written and spoken descriptions of the WAB Picnic scene, based on a pre-defined CU corpus. We then calculated the ratio of CU to U (CU/U Ratio) as a measure of content density. Our cohort included 115 participants (20 control participants for written, 20 control participants for spoken, 28 participants with nfvPPA, 30 with lvPPA, and 17 with svPPA). We compared written language between patients with PPA and control participants and written to spoken language in patients with the three variants of PPA. Finally, we analyzed CU and U in relation to the Progressive Aphasia Severity Scale Sum of Boxes and the Clinical Dementia Rating Sum of Boxes. Our program identified CU with a validity of 99.7% (95%CI 99.5 to 99.8) compared to manual annotation of the samples. All patients with PPA wrote fewer total units than controls (*p<*0.001). Patients with lvPPA (*p=*0.013) and svPPA (0.004) wrote fewer CU than controls. The CU/U Ratio was higher in nfvPPA and svPPA than controls (*p=*0.019 in both cases), but no different between lvPPA patients and controls (*p=*0.962). Participants with lvPPA (*p<*0.001) and svPPA (*p=*0.04) produced fewer CU in written samples compared to spoken. A two-way ANOVA showed all groups produced fewer units in written samples compared to spoken (*p<*0.001). However, the decrease in written CU compared to spoken was smaller than the decrease in written units compared to spoken in participants with PPA, resulting in a larger written CU/U Ratio when compared to spoken language (*p<*0.001). nfvPPA patients produced correlated written and spoken CU (*R=*0.5, *p=*0.009) and total units (*R=*0.64, *p<*0.001), but this was not the case for lvPPA or svPPA. Considering all PPA patients, fewer CU were produced in those with greater aphasia severity (PASS SoB, R=-0.24, p=0.04) and dementia severity (CDR SoB, R=-0.34, p=0.004). In conclusion, we observed reduced written content in patients with PPA compared to controls, with a preference for content over non-content units in patients with nfvPPA and svPPA. When comparing written to spoken language, we observed a similar “telegraphic” style in both modalities in patients with nfvPPA, which was different from patients with svPPA and lvPPA, who use significantly less non-content units in writing than in speech. Lastly, we show how our program provides a time-efficient tool, which could enable feedback and tracking of writing as an important feature of language and cognition.

## Introduction

Primary progressive aphasia (PPA) is a clinical syndrome in which aphasia is the initial predominant symptom, usually resulting from Frontotemporal Lobar Degeneration or Alzheimer’s disease.^1^ The characteristics of aphasia in PPA are heterogeneous, with many patients presenting with a profile of language impairments that can be classified into one of three subtypes: the non-fluent/agrammatic variant (nfvPPA), the logopenic variant (lvPPA), or the semantic variant (svPPA).^2^ Although the characterization and classification of PPA patients’ aphasia is primarily done based on clinical history taken from the patient and an informant, interview of the patient, and formal assessment of spoken language, in our experience, patients commonly have written language abnormalities as well, and some have predominant alexia^3,4^ or agraphia.^5^ For the three major PPA variants, the only written language characteristic that is part of the diagnostic criteria is surface dyslexia or dysgraphia in svPPA.^2^ Despite the important role of written language in contemporary everyday life (especially in societies dominated by email, online posts, and text message communication), abnormalities in functional written communication have been sparsely investigated in PPA. Yet because some patients may be able to communicate more effectively through some form of writing or typing than through speech, the characterization and measurement of strengths and weaknesses in written communication should be part of the assessment of patients with PPA. Ultimately, the comparison of spoken and written language content in a patient with PPA may offer clinicians opportunities to develop compensatory strategies to maximize functional communication.

Most studies of writing impairments in PPA have focused on spelling; very few have investigated functional communication. In a comparison of written picture descriptions in patients with nfvPPA to patients with progressive supranuclear palsy (PSP) and controls, patients with nfvPPA have reduced length, speed, and information units compared to controls,^6^ as well as reduced number of written words compared to PSP.^7^ Long term follow-up of written language in one patient with nfvPPA also showed a decrease in written word output and amount of information, as well as a decrease in sentence complexity over time and increased dependence on nouns over verbs.^8^ A study comparing written picture descriptions of patients with lvPPA to those of amnestic mild cognitive impairment (MCI) and dementia likely due to Alzheimer Disease reported that patients with lvPPA had increased letter insertion errors and a higher verb use.^9^ A case study of a patient with an unspecified type of PPA and atrophy in the left inferior frontal gyrus showed difficulty in written word construction to convey meaning.^10^ There have been two case reports on progressive changes in writing in patients with svPPA, displaying an increase in written output, including writing books or increased diary entries, though with a decrease in complexity over time.^11,12^ When comparing written to spoken spelling in a mixed group of participants with PPA, Henry, *et al*.^13^ showed parallel changes in both modalities, suggesting proportional impairments deriving from core linguistic dysfunction in phonology and semantics. However, in a study of agrammatism in patients with nfvPPA compared to patients with Primary Progressive Apraxia of Speech (PPAOS), the nfvPPA group made more grammatical errors in speech than in writing relative to the PPAOS group.^14^

In this study, our goals—using a language-elicitation test (WAB picnic scene)—were to compare the content and quantity of written communication between the three PPA variants and cognitively normal control participants, compare written to spoken language in PPA patients, and examine the relationships of written language output to the severity of aphasia and the severity of overall cognitive impairment. We developed an automated program which uses a language parser and quantifies content units (CU) and total units (words and word attempts) in transcribed spoken and written language samples, employing a pre-defined CU corpus.^15^ Based on our prior study of spoken language,^15^ we hypothesized a decrease in the total number of written CU and units in participants with PPA compared to controls. In cases where syntax may be simplified, such as in participants with nfvPPA, we expected an increase in the CU to unit ratio. Prior studies comparing written to spoken language in healthy individuals (from primary school to graduate students), have shown a relative decrease in written output with a relative increase in the density of CU in written language compared to spoken.^16^ Therefore, we hypothesized an overall decrease in written compared to spoken CU in both participants with PPA and controls. Based on our clinical experience, we expected fewer task-irrelevant units, such as self-referential and tangential language, in written than in spoken language, especially in lvPPA and svPPA. Also based on our clinical experience, writing can sometimes be less impaired than speech in nfvPPA (especially those with concomitant motor speech disorders); in this case, the amount of CU in writing and in speech may be similar. Finally, we expected a proportional decrease in content in patients with relatively greater aphasia severity (Progressive Aphasia Severity Scale Sum of Boxes) and in those with greater cognitive and functional impairment (Clinical Dementia Rating Sum of Boxes).

## Materials and methods

### Participants

Seventy-five individuals diagnosed with PPA were included in this study, all of whom were recruited through the Massachusetts General Hospital (MGH) Frontotemporal Disorders Unit PPA program. All participants (and their care partners for patients with PPA) gave written informed consent in accordance with guidelines established by the Mass General Brigham Healthcare System Institutional Review Boards which govern human subjects research at Massachusetts General Hospital. See **Table 1** for full demographic and clinical data. All patients received a standard clinical evaluation comprising a structured history obtained from both patient and informant, comprehensive medical, neurological, and psychiatric history and exams, neuropsychological and speech-language assessments, and a clinical brain MRI that was visually inspected for 1) regional atrophy consistent with or not consistent with a given syndromic diagnosis, and 2) other focal brain lesions or evidence of cerebrovascular disease. Clinical formulation was performed through consensus conference by our multidisciplinary team of neurologists, psychiatrists, neuropsychologists, and speech and language pathologists.^17^ All patients included in this study met the diagnostic criteria for PPA and all were able to be subclassified into one of the major subtypes with a clinical imaging-supported atrophy pattern: the non-fluent/agrammatic variant (nfvPPA), semantic variant (svPPA), or logopenic variant (lvPPA) of PPA.^2^ Furthermore, no patient had any other focal brain lesions or significant cerebrovascular disease (e.g., previous strokes, cerebral hemorrhages, meningiomas); none had major psychiatric illness not adequately treated; and all were native speakers of English. From the structured evaluations, each patient had a Clinical Dementia Rating (CDR) global score, a CDR Sum-of-Boxes (CDR-SoB) score, a CDR Supplemental Language Box score, and a Progressive Aphasia Severity Scale Sum-of-Boxes (PASS-SoB) score.^18^ The PASS is a clinical instrument used to rate presence and severity of impairment in specific domains of speech and language.^18^ The CDR is a clinical dementia rating tool which reflects general aspects of cognition and activities of daily living.^19^ The cohort analyzed here included 28 patients with nfvPPA, 30 patients with lvPPA, and 17 patients with svPPA. Patients with mixed forms of aphasia or primary progressive apraxia of speech were excluded from the analysis. Three patients with nfvPPA and three patients with lvPPA were not able to complete the written task and were therefore not included in this analysis despite their ability to produce spoken samples.

**Table 1.** Research participant demographic data.

|  | **Controls** | **nfvPPA** | **lvPPA** | **svPPA** | **p** |
| --- | --- | --- | --- | --- | --- |
| **Sample size (n)** | 20/20<sup>1</sup> | 28 | 30 | 17 | NA |
| **Age** | 61.08±10.64 | 71.21±7.78 | 69.83±7 | 66.82±7.74 | <0.001* |
| **Sex (female)** | 57.5% | 57.1% | 46.6% | 58.8% | 0.779 |
| **Years of education** | 15.85±1.34 | 15.8±2.95 | 16.52±2.44 | 16.44±2.48 | 0.478 |
| **Handedness (right)** | 87.5% | 85.7% | 76% | 88.2% | 0.057 |
| **Years from symptom onset** | NA | 3.79±2.06 | 4.5±2.9 | 4.56±1.12 | 0.458 |
| **CDR CGS** | NA | 0.404±0.28 | 0.450±0.24 | 0.531±0.22 | 0.291 |
| **CDR SoB** | NA | 5.90±4.35 | 5.48±2.09 | 4.68±1.86 | 0.451 |
| **CDR Language** | NA | 0.821±0.53 | 0.77±0.35 | 0.74±0.26 | 0.777 |
| **PASS SoB** | NA | 5.90±4.35 | 5.48±2.09 | 4.68±1.86 | 0.451 |

<sup>1</sup> 20 control participants for written language, and 20 control participants for spoken language. Data in that column corresponds to total (40) control participants
\*nfvPPA and lvPPA relative to controls

Spoken samples were obtained from 20 age-matched healthy controls – with no self-reported history of neurologic or psychiatric disorders – from the Speech and Feeding Disorders Laboratory at MGH Institute of Health Professions. Written samples from 20 age-matched controls were obtained from the Amazon’s Mechanical Turk volunteers after they confirmed lack of any neurological or language related abnormalities. Demographic features from these participants are presented in Table 1.

### Sample collection

Participants performed spoken and written descriptions of the Western Aphasia Battery – Revised (WAB-R) ‘Picnic Scene’ task. Participants were instructed to produce both spoken and written descriptions using full sentences. Spoken samples were transcribed with the aid of Microsoft Word Version 16.57 dictation software, and manually corrected by either N.R. or S.J.H. Handwritten samples from patients were manually transcribed to text files by S.J.H, A.J, or I.H. (see acknowledgements). Spelling errors were manually corrected upon transcription and quantified separately. Written samples from controls were typed and did not require any additional text processing.

### Definitions

Quantified terms are defined as follows:

- Content unit (CU): “Correct information units are words that are intelligible in context, accurate in relation to the picture or topic, and relevant to and informative about the content of the picture or the topic. Words do not have to be used in a grammatically correct manner to be included in the correct information count.”^20^ Following Berube *et al*.^*21*^ the dictionary of CU utilized was compiled from CUs that were each mentioned in spoken descriptions by at least three healthy controls. The 64-CU dictionary used was previously published by Gallée *et al*.^15^ Each CU is only counted once, regardless of how many times it is mentioned in a sample. The previously published CU dictionary^15^ also grouped morphological variants within one single CU. For example, the nouns “girl” and “daughter” are listed as CU #12. Therefore, if one participant used both words (girl and daughter), they would only be counted as one CU. We followed the same dictionary for consistency.
- Unambiguous CU: Unambiguously refer to a particular object/entity, action, or property of a specific object/entity in the picture^15^. The term used can only apply to a single feature within the scene. For example, there is only one dog in the scene, dog is an unambiguous unit.
- Ambiguous CU: Do not unambiguously refer to a particular aspect of the picture.^15^ The term in isolation can correspond to more than one feature within the scene. The term will be labeled as ambiguous even if context allows to make a confident estimate of a feature in the scene. For example, both the dog and the boy are running, the term ‘running’ is an ambiguous CU.
- Self-referential: Any first-person pronoun – I, we, us.
- Unit of speech (unit; i.e., U): Total count of every word, non-word, false start.^15^ Contractions are counted as two units, for example, “they’re” = 2 units.
- CU/Unit Ratio (CU/U Ratio): Total CU (repeated CU only counted once) divided by total units. In other publications this corresponds to the term “informativeness”.^15^ However, the term informativeness may be misleading since a higher value does not always reflect a language sample that conveys greater meaning, as it not only depends on the number of CU (numerator) but also the total number of units (denominator). For example, a list of CU without basic sentence structures will have a higher CU/U Ratio but convey less meaning because the words that capture relationships between CU are not present.

### Program

We used Quantitext, a text analysis toolbox we developed in the Frontotemporal Disorders Unit of MGH, to automatically produce a set of quantitative language metrics. The goal of developing this package is to increase the precision and objectivity of language assessments while reducing human labor.^22^ The toolbox uses a number of natural language processing toolkits and software such as Stanford Parser,^23^ spaCy,^24^ as well as text analysis libraries in R. Quantitext receives transcribed language samples as input and generates as outputs a number of metrics such as sentence length, log word frequency, log syntax frequency, content units, total units, efficiency of lexical and syntactic items, and part of speech tags. To specify content units, the toolbox first generates a python dictionary using a predefined set of words as previously described^15^ and then uses this dictionary to automatically identify all content units in new texts that it receives as input. The program then counts the content units and all units in the language sample of each participant. To measure the program’s validity, each sample was then manually annotated to check for omissions and proper counting.

### Statistical analysis

Statistical analysis performed with IBM SPSS Statistics Version 28.0.0.0 and Prism 9 for macOS Version 9.3.0 (345). Our program’s validity was assessed with a Pearson bivariate correlation analysis. Ordinal and nominal values were compared between groups using Chi square. Normally distributed values were compared between groups using a one-way ANOVA. When significant differences were present, post-hoc analysis was performed with Tukey post-hoc multiple comparisons procedure. Within spoken to written and between group comparisons were performed with a two-way ANOVA. When comparing written to spoken language between the three main PPA subtypes, a one-between (PPA subtypes) – one-within (written vs spoken) ANOVA was used. Within participant analysis was done exclusively for written and spoken samples collected within 6 months of each other. Bivariate Pearson correlation analysis was performed when comparing language measurements to clinical scales. Results were considered statistically significant when *p<*0.05, and trends are reported when *p<*0.1.

### Data availability

The code for the program described in this manuscript is available in the public repository. The data that support the findings of this study are available on request from the corresponding author. The data are not publicly available due to the presence of information that could compromise the privacy of research participants. Derived data supporting the findings of this study are available from the corresponding author on request.

## Results

### Participants

The clinical and demographic characteristics of the participants are shown in Table 1. Participants with nfvPPA and lvPPA were older in age when compared to control participants. Otherwise, there were no significant differences between control participants and participants with the three different PPA subtypes in sex distribution, years of education, and handedness. There were no significant differences between the three PPA subtypes in years from symptom onset, average CDR scores, CDR SoB, CDR Supplemental Language Box, and PASS SoB.

### Program validity

The automated program identifies CUs with a validity of 99.7% (CI 99.5– 99.8, p<0.001) compared to manual annotation of the samples.

### Functional written communication in PPA

As expected and shown in Figure 1 and Table 2, all patients with PPA wrote fewer total units than controls (*F*(3, 91)=[7.778], *p<*0.001), and there were no differences between the three PPA variant groups. As for CUs, patients with lvPPA and svPPA wrote fewer than controls (one-way ANOVA *F*(3, 91)=[4.897], p=0.003; Tukey’s post-hoc tests *p=*0.013 for lvPPA and 0.004 for svPPA). The average number of CUs written by the nfvPPA group was lower than controls, but this was not a statistically significant difference (Tukey’s post-hoc test *p*=0.14). There were no differences between the three PPA variant groups in written CUs. The CU/U Ratio was higher in nfvPPA and svPPA than controls (*F*(3, 91)=[5.592], *p=*0.019), but no different between lvPPA patients and controls (*p=*0.962). The CU/U Ratio was higher in nfvPPA and svPPA than lvPPA (Tukey’s post-hoc test *p*=0.031 for nfvPPA and p=0.033 for svPPA).

**Table 2.** Measurements of Written Language.

| **Group** | **CU** | **Units** | **CU/U Ratio** | **n** |
| --- | --- | --- | --- | --- |
| Control | 18.5±9.32 | 61.70±27.86 | 0.30±0.05 | 20 |
| All PPA | 12.24±6.74 | 34.68±21.63 | 0.38±0.16 | 75 |
| nfvPPA | 13.89±6.33 | 34.64±18.87 | 0.42±0.14 | 28 |
| lvPPA | 11.97±7.87 | 38.00±24.54 | 0.32±0.12 | 30 |
| svPPA | 10.00±4.50 | 28.88±20.36 | 0.44±0.22 | 17 |

**Figure 1.**
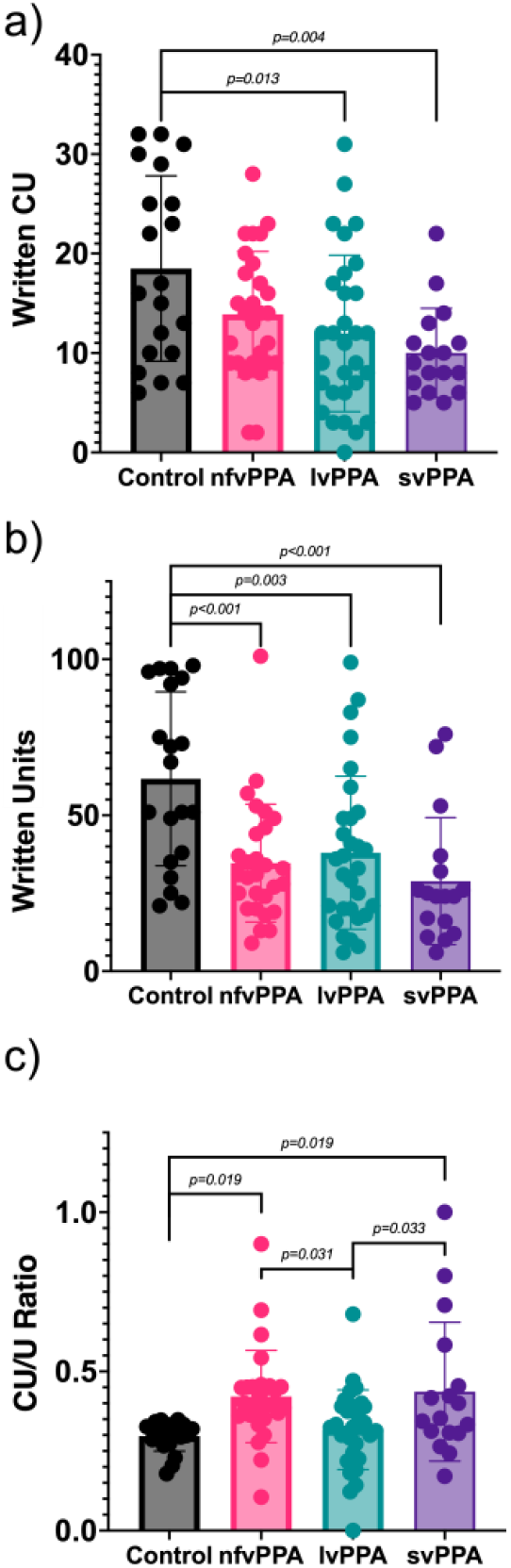
Comparison of written language between participants with PPA and controls. Graphs illustrate written Content Units (CUs) **(a)**, total Units (U) **(b)**, CU/U Ratio **(c)** in controls and the three PPA variants. Error bars indicate one standard deviation. Statistically significant differences between pairs of groups are indicated by lines.

We obtained the mean and standard deviation for the written CU/U ratio in all samples (0.364±0.151) and qualitatively examined the samples of patients with CU/U Ratios one standard deviation below (0.213) and one standard deviation above the mean (0.516). There were nine participants with a CU/U ratio at least one standard deviation below the mean. Those nine participants comprised two controls, one patient with nfvPPA, one with svPPA, and five with lvPPA. These samples either had a very small number of CUs, or had a larger number of tangential, vague, and self-referential units which did not add content to the description. There were also nine participants with a CU/U ratio at least one standard deviation above the mean. Those nine participants comprised four patients with nfvPPA, four with svPPA, and one with lvPPA. These descriptions tended to be short, and in some cases consisted of a list of CUs with a very simple or absent sentence structure. For example: “Woman pouring drink” all correspond to CU.

### Comparison of written and spoken language in PPA patients

Figure 2 and Table 3 show comparisons of written versus spoken language in the three PPA subtypes. Examining each of the three groups of PPA variant, the overall number of CUs was smaller in written versus spoken language (*F*(1,67)=[22.62], *p<*0.001). Tukey’s post-hoc tests showed that participants with lvPPA (*p<*0.001) and svPPA (*p=*0.044) produced a lower number of written CUs versus spoken CUs, but this effect was only present as a trend in participants with nfvPPA (*p=*0.09). The overall number of total units was smaller in written versus spoken language in all subtypes (*F*(1, 67)=[96.75], *p<*0.001). The CU/U Ratio was higher in written than spoken language for all PPA subtypes (*F*(1, 67)=[96.49], *p<*0.001).

**Table 3.**
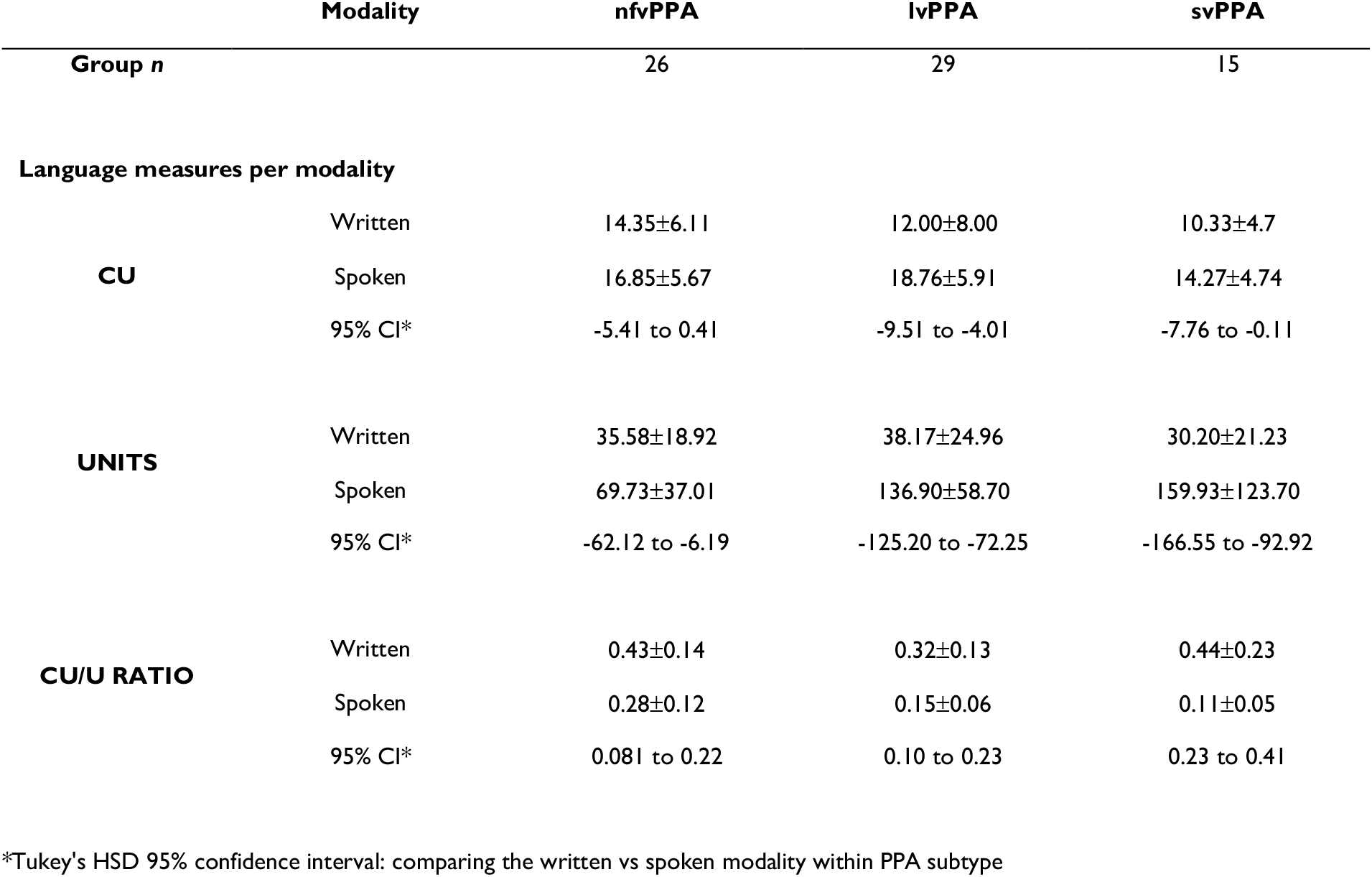
Comparison in language measures according to modality in the three main PPA subtypes.

**Figure 2.**
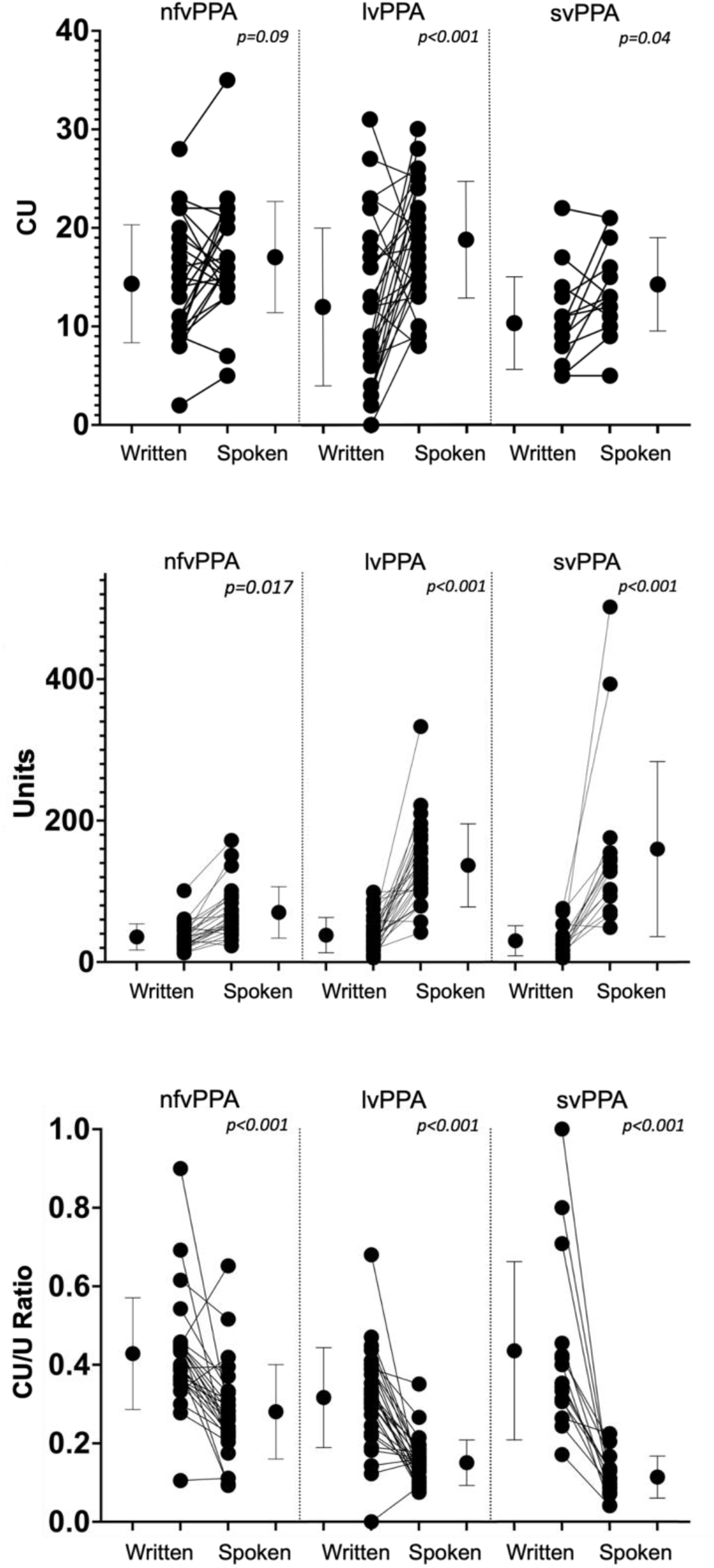
Comparison of written and spoken language within PPA participants. Each graph shows the mean and one standard deviation for the group of participants on the left (written) and right (spoken) of each panel, with individual datapoints from the same participant connected by a line in the middle. Data from each of the PPA variants are displayed separately. The rows of graphs show Content Units (CUs) **(a)**, total Units **(b)**, and the CU/U Ratio **(c)**. The *p* values at the top of each panel correspond to Tukey HSD post-hoc contrasts, in a two-way one-between/one-within ANOVA comparing the written to spoken modality within each PPA subtype.

At the individual participant level, although the total number of total units was smaller in written compared to spoken language in nearly every single patient with PPA (92.86% of participants), this was not so universally true for CUs. Written CUs were lower than spoken CUs in 61.54% of participants with nfvPPA, 79.31% of those with lvPPA, and 80.00% of those with svPPA. The CU/U Ratio was higher in written than spoken language for 84.62% of participants with nfvPPA, 89.66% of those with lvPPA, and 100% of those with svPPA. The differences in these values, illustrated in Figure 2, reflect the intra-group variability in written vs. spoken language production within and between the different PPA subtypes.

As illustrated in Figure 3, we performed correlation analysis between the spoken and written modality for CU, total units, and the CU/U Ratio in each PPA variant. In the nfvPPA group only, there is a correlation between the number of spoken CU and written CU (*R=*0.5, *p=*0.0086) and between the number of spoken and written total units (*R=*0.64, *p<*0.001). There is a trend toward a correlation between spoken and written CU in svPPA (*R=*0.4, *p=*0.13). This was not the case for written vs spoken CU (R=0.12, p=0.54) and units (R=0.08, p=0.66) in lvPPA, or units in svPPA (R=0.27, p=0.33). There was also no correlation between spoken and written CU/U Ratio in any of the subtypes (all *p* values > 0.24).

**Figure 3.**
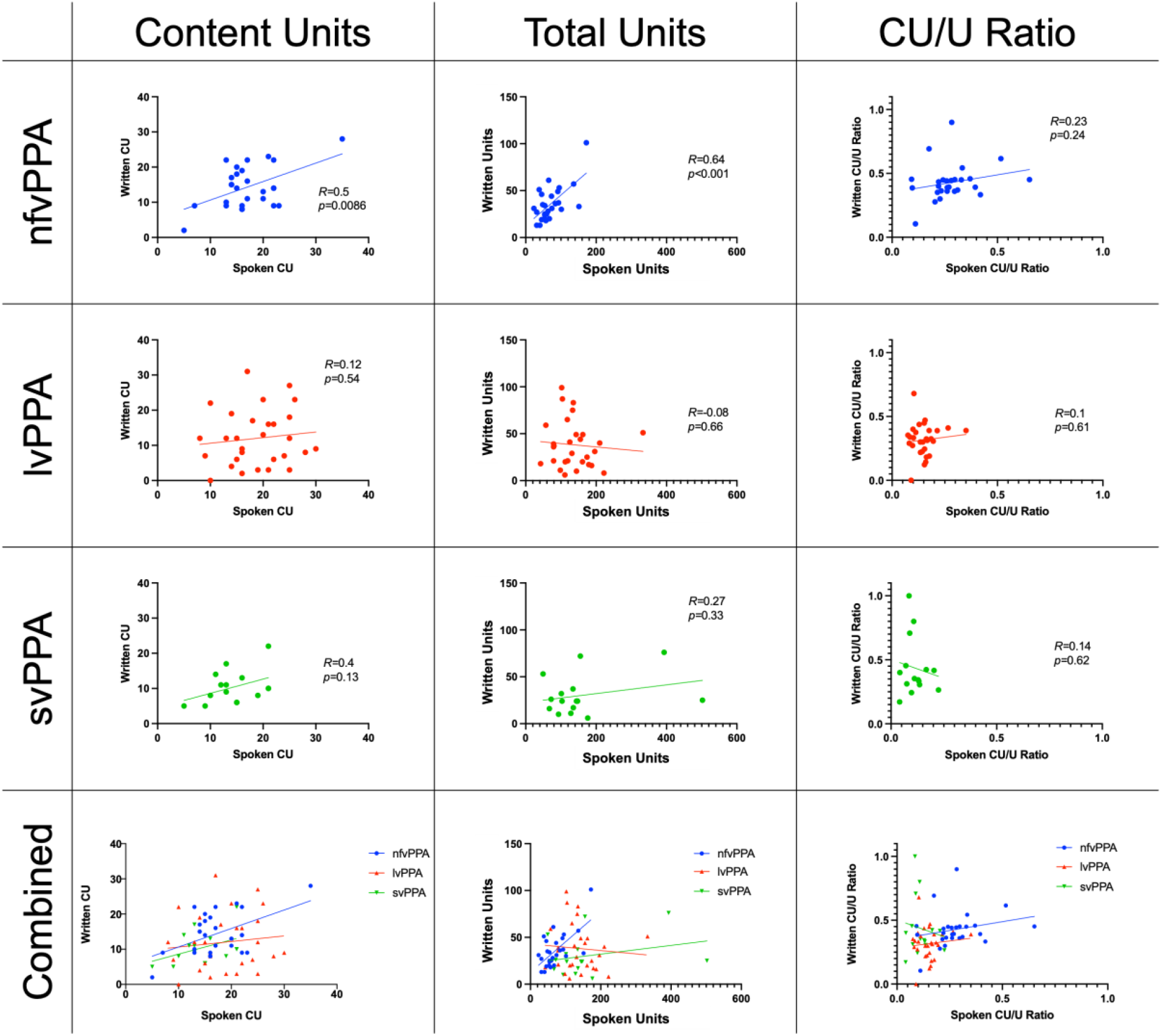
Scatterplots comparing spoken and written CU, total Units and CU/U Ratio. In all graphs, written language is on the Y axis and spoken language is on the X axis. The left column shows CU, the middle column shows total Units, and the right column shows CU/U Ratio. The rows separate the three PPA variants, with the bottom row showing the three variants combined. Statistics included refer to linear correlation analysis.

The scatterplots show several features worth highlighting: (1) in the case of spoken vs written CU, especially in the case of lvPPA, there are some participants who say many more CU than they write, and vice-versa; (2) for the CU/U Ratio, svPPA patients exhibit an extreme distribution with an almost vertical plane, reflecting much greater variability in written CU/U Ratio compared to spoken CU/U Ratio; lvPPA patients show a somewhat similar but less extreme effect. In both cases, the pattern appears to be due to the relative absence of empty language in the written modality. This is in contrast with patients with nfvPPA, where participants have a relatively higher CU/U Ratio in both writing and speech.

We also examined the ambiguity of CUs, comparing written to spoken language samples (Figure 4). In the spoken modality, there were significant differences in the proportion of unambiguous and ambiguous CU to total CU, specifically between patients with nfvPPA and controls, as well as between nfvPPA and the other two PPA subtypes (one-way ANOVA *F*(3, 94)=[10.281], *p<*0.001), with a preference towards unambiguous CU. As seen in Figure 4, though not statistically significant, patients with svPPA have a relative predilection for ambiguous CU (relatively fewer unambiguous CU). Despite a similar general pattern observed, when compared to the spoken modality, there were no significant differences in the proportion of written unambiguous and ambiguous CU to total CU between PPA and controls, or between the different PPA subtypes (one-way ANOVA *F*(3, 94)=[1.197], *p=*0.316). This lack of significant differences may be explained by increased variability in the written modality when compared to the spoken modality. A three-between groups ANOVA showed there were no differences in the distribution of ambiguous and unambiguous CU between the three PPA subtypes and controls, when comparing the written to the spoken modality (*F*(3, 342)=[0.54], *p=*0.65).

**Figure 4.**
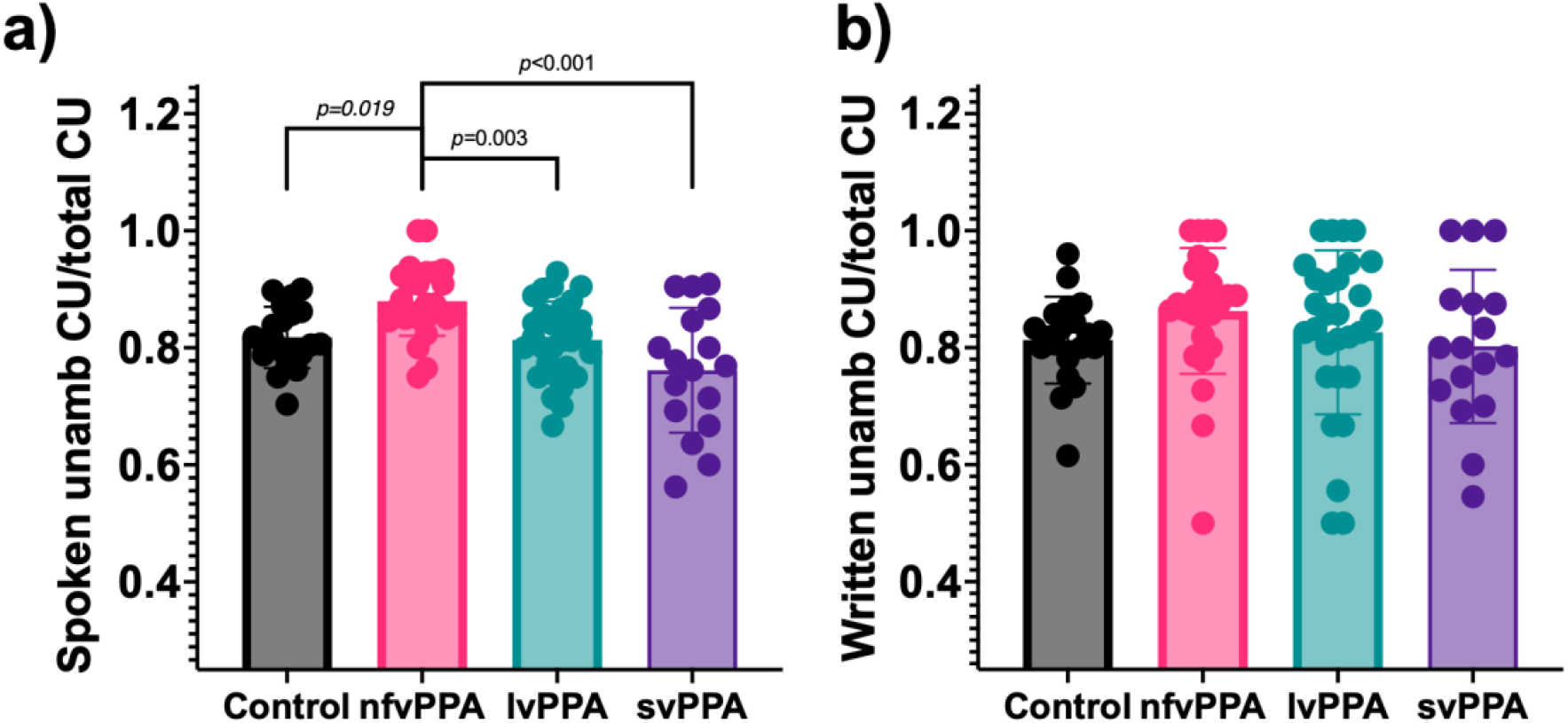
Spoken and written unambiguous CUs in control participants and the three PPA subtypes. Graphs depict the average proportion of CUs that are ambiguous relative to total CU in spoken **(a)** and written **(b)** language, in control participants and the three PPA variants. Statistically significant differences between pairs of groups are indicated by lines. Ambiguous CU correspond to the remaining CU within the total (i.e., unambiguous CU/total CU + ambiguous CU/total CU = 1).

Finally, based on our prior observations of self-referential speech in svPPA,^15^ we examined self-referential pronouns in writing. There were no differences in written self-referential pronouns between the three PPA subtypes (nfvPPA 0.07+/-0.38; lvPPA 0.27+/- 0.78; svPPA 0.00) and controls (0.15+/-0.37) (*F*(3, 91)=[1.184], *p=*0.32). Participants with svPPA had more spoken self-referential pronouns (7.29+/-9.74) than controls or participants with nfvPPA or lvPPA (1.55+/-1.50, 0.96*+/-*2.33, and 2.80+/-2.71 respectively) (*F*(3, 91)=[7.408], *p<*0.001).

### Relationships between language measurements and severity of clinical impairment in PPA

Although the patients in this sample had mild-to-moderate degrees of aphasia and only prodromal to mild dementia, we examined relationships between written language metrics of interest and aphasia severity (PASS Sum of Boxes) and dementia severity (CDR Sum of Boxes). As seen in Figure 5, correlation analysis showed small effect size relationships indicating that written CUs are reduced in patients with greater dementia severity (CDR SoB *R=-*0.34, *p=*0.004) and greater aphasia severity (PASS SoB *R=-*0.24, *p=*0.04). A reduction in spoken CU was also observed with greater dementia severity (CDR SoB *R=-*0.31, *p=*0.011) with a trend toward the same relationship with greater PASS SoB score (*R=-*0.21, *p=*0.078). Similarly, for total written units a decrease was observed with increasing CDR SoB (*R=-*0.30, *p=*0.012) and a trend with PASS SoB (*R=-*0.22, *p=*0.067). No correlation was observed between spoken total units and CDR SoB (*R=*0.08, *p=*0.497) or PASS SoB (*R=*0.07, *p=*0.561). Regarding the CU/U Ratio, a correlation was only observed between spoken CU/U Ratio and CDR SoB (*R=*0.29, *p=*0.018), but not with PASS SoB (*R=*0.08, *p=*0.527) or between written CU/U Ratio and CDR SoB (*R=*0.03, *p=*0.753) or PASS SoB (*R=*0.04, *p=*0.718). This study was not powered to examine these relationships in each PPA variant.

**Figure 5.**
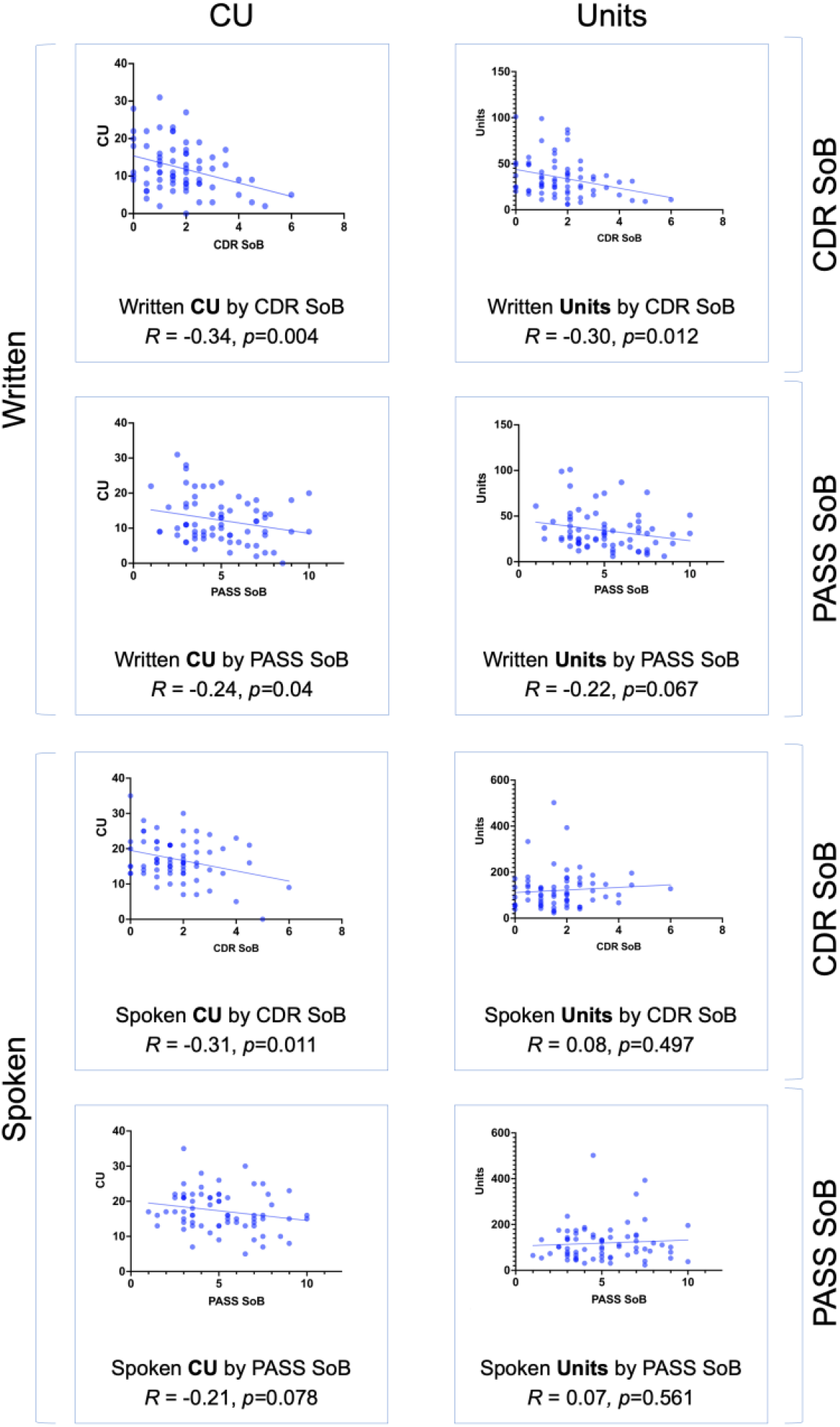
Correlation analysis of written and spoken CU and total units from all participants with PPA against their respective clinical rating scores. The left column corresponds to CU and the right to total units. The top four boxes correspond to the written modality and the bottom four to spoken. Each modality is further subdivided into correlation with CDR SoB or PASS SoB as shown on the right. Statistics shown correspond to linear correlation analysis.

## Discussion

In this work, we investigated the content of picture-elicited written language samples in PPA patients and healthy controls using an automated computational analytic method. We also compared the content of written language samples to spoken language samples elicited using the same task. We showed that despite a decrease in written CUs and units in participants with PPA, there was a relative increase in the density of content in written language compared to spoken language, especially in patients with lvPPA and svPPA due to a lesser amount of “empty” language. In patients with nfvPPA, written language was frequently “telegraphic” as previously described. We observed similar reductions in the generation of ambiguous CUs between writing and speech in nfvPPA. Finally, we showed that CUs decrease proportionally to greater severity of aphasia or cognitive/functional impairment.

In the analysis of written language in PPA, spelling has been relatively thoroughly explored,^25-29^ but other elements of written language have received very little attention. Written language in nfvPPA has been described as “telegraphic”^6,30^ which, as hypothesized, we found here as well: nfvPPA patients prefer content words over non-content words, ultimately leading to a higher CU/U Ratio. In contrast, Graham, *et al*.^*6*^ did not find higher ‘information units per word’ in patients with nfvPPA compared to controls, which may be due to controls in our study writing or saying fewer CU. This in turn could be due to a difference in instructions, our “describe this picture using full sentences” as opposed to Graham’s ‘‘Tell/write everything you see going on in this picture’’. Lastly, we extend prior work to also demonstrate a similar increase in written CU density in svPPA, when compared to controls, and in all PPA subtypes when comparing writing to the patients’ own spoken language.

The “primary systems” hypothesis predicts that parallel changes are expected to occur in spoken and written language.^31^ This hypothesis has been supported in spelling in PPA,^13^ but to our knowledge not using other measures of written vs. spoken language. We observed similarities in the pattern of ambiguous and unambiguous CU in both written and spoken language in all PPA variants, again supportive of the primary systems hypothesis. However, when comparing CUs, units, and CU density (CU/U Ratio), we observed a disproportionate decrease in total units relative to CU between written and spoken samples, predominantly in lvPPA and svPPA but also in nfvPPA, leading to a relative increase in the CU/U Ratio. A similar increase in ‘information units per word’ in written compared to spoken descriptions had been previously reported by Graham, *et al* in patients with nfvPPA and controls.^6^ The relative sparing of written language in nfvPPA, when compared to the other PPA subtypes, could account for the milder decrease in CU in writing compared to speech.

This difference in written versus spoken language has not been previously published in lvPPA or svPPA. Given that Alzheimer’s disease pathology is most often the underlying etiology for lvPPA, and their shared neuroanatomic distribution affecting the temporo-parietal junction,^2,32^ previous reports on writing in lvPPA have provided comparisons with patients with MCI or Alzheimer’s disease dementia, demonstrating higher verb use in patients with lvPPA.^9^ Also, studies comparing written to spoken language in Alzheimer’s disease dementia have shown differences in the information pattern between both modalities, with written descriptions being as informative, but shorter and syntactically simplified compared to spoken descriptions.^33^ The later finding has some overlap with our own, in that written descriptions were shorter and simplified compared to spoken in patients with lvPPA. However, CU were fewer in the written than the spoken modality, which may argue against equivalent informativeness despite the lack of significant empty language in writing compared to speech.

Studies of writing in svPPA include reports of increased creativity in writing in three patients with svPPA^34^, as well as a longitudinal writing analysis in two patients with svPPA^11,12^. Hypergraphia in these cases may be attributed to behavioral changes commonly observed in svPPA^2^. Heitkamp, *et al*.^11^ studied longitudinally the personal diary of a patient with svPPA, reporting an increase in the length of the diary entries with a concomitant decrease in vocabulary over time. Also, they noticed an increase in ambiguity (which we observed in participants with svPPA as well), a decrease in the variety of vocabulary, an increase in type-token ratio (which may be comparable to the increase in CU/U Ratio we report here), and simplified syntax at later stages.^11^ In the second longitudinal analysis of written text in a patient with svPPA, Hwang, *et al*.^12^ also showed a decrease in lexical sophistication and an increase in ambiguity, the latter similar to our findings.

One goal of spoken or written descriptions is to convey as much information as efficiently as possible, even if a language or other cognitive impairment interferes with the process. Ravid, *et al*.^16^, showed that information density depends on the language modality, with significantly more ancillary (non-descriptive) material found in spoken when compared to written narratives. These findings may reflect more rapid “online” processing in spoken narratives. In written narratives, there may be a higher cognitive demand, but also more planning and monitoring (“offline production”).^16^ The manner in which a patient with PPA maximizes their ability to convey information is expected to depend on the nature of their language impairment and the language modality being employed. For example, given that participants with PPA have an overall decreased output in written language, they may prioritize words that will be more informative, at the expense of ancillary material and repairs.^16^ We recently reported this type of trade-off between word complexity and syntax in spoken language in PPA.^35^ In spoken language in the case of lvPPA and svPPA, specific word retrieval is challenged despite relatively preserved fluency in conversation. Participants with these subtypes may therefore opt to say more overall, including circumlocutions and repairs, with the overall objective of conveying as much information as possible, at the expense of a decrease in information density. The increase in CU/U Ratios in written compared to spoken language may also reflect the optimization of investing a higher cognitive effort in fewer yet more informative language units.

It is important to explore the difference between the term informativeness and the CU/U Ratio. The term informativeness has been used to describe the communication of CUs relative to all words in speech^15,21^, and calculated with the same equation we used here as the CU/U Ratio. We chose not to use the term informativeness here because, even in patients with relatively mild PPA, some individuals simply wrote a list of CUs without full sentence structure (despite the task instructions to write in full sentences). This occurred with lower frequency in speech samples, and usually in more impaired patients. As such, the higher CU/U ratio reflected the high percentage of content words. While a list of nouns or verbs does provide content, it lacks informativeness about the relationship between the items. We found this to be the case only with some of the more severely impaired patients, whose writing samples contain few or no grammatical words, or whose samples are very short. Future investigation would be helpful to determine if the rating system presented here has the greatest utility in patients at a mild to moderate severity level of impairment, and less so for more severely impaired patients. Nevertheless, the CU/U Ratio helps to identify outliers whose language can be further examined quantitatively or qualitatively, as we explored in our results.

To the best of our knowledge, there is not software that is equivalent to the code described in this manuscript. Clarke *et al*.^36^ thoroughly explored the multiple technologies that have been developed for automated language analysis and their role in mild cognitive impairment, Alzheimer’s disease, and dementia with Lewy bodies. However, such technologies have scarcely been tested in PPA. Two such examples include the automated analysis of spelling errors^37^ and the automated analysis of two written works in a patient with svPPA.^12^ In-depth analysis of written language can be time-consuming, and therefore is not routinely incorporated into clinical practice. As many societies become more familiarized with technologies, from typing on a smartphone to typing on a computer, tools that analyze written language can rapidly provide objective and likely clinically relevant information.

In this work, we also determined whether the reduction in written and spoken CU and total units were merely a reflection of general cognition in participants with PPA. The mild-to-moderate correlation observed between the CDR SoB and written CU and units, as well as spoken CU, shows how these measures are only a partial reflection of general cognition, though may act as an indirect marker. Interestingly, Mazzeo, *et al*.^38^ recently showed how written phrases under dictation can act as a predictor for total loss of speech in AD-related PPA.

The main limitation of this study is the fact that two separate control groups were used, one for spoken samples and one for writing. This prevented a within-group analysis comparing language modalities from being performed in the control group. Furthermore, the control group for written samples performed typed descriptions of the picnic scene, as opposed to the handwritten descriptions by participants with PPA. Based on time and effort required to type compared to handwrite, we would expect typed descriptions to be longer than handwritten descriptions. This may accentuate the difference in the total CU and units when comparing control participants to participants with PPA; however, the reduction observed in the participants with PPA is still more likely to be due to word retrieval difficulties. Also, a comparable CU/U Ratio between the written and spoken samples in the control groups further supports the validity of the data as representations of healthy human language production. Another limitation is the high level of education in our samples, which may limit the generalizability of our findings. Finally, the correlation analysis with clinical rating scales was performed by pooling the samples of all participants with PPA, since the study was not powered to analyze these variables within PPA variants separately.

There is much more to investigate in the writing of patients with PPA. Future studies may include analyses of specific elements of written language, such as grammar, word familiarity, syntax, verb and noun proportions, and their comparison to speech. Future studies should examine language longitudinally. We would expect to find stronger relationships between written and spoken CU if they were tracked over time within individual patients.^8,11,12,39^ Future studies may also examine naturalistic production of written language, such as analyzing text messages or emails, which may more closely reflect a patient’s day-to-day functional communication. Lastly, it would be valuable to examine the neural correlates of written language impairments using neuroimaging.

## Data Availability

The code for the program described in this manuscript will be available in a public repository. The data that support the findings of this study are available on request from the corresponding author. The data are not publicly available due to the presence of information that could compromise the privacy of research participants. Derived data supporting the findings of this study are available from the corresponding author on request.

## Abbreviations

CDR SoB: Clinical Dementia Rating Sum of Boxes
CI: confidence interval
CU: content unit(s)
CU/U Ratio: Content Unit/Unit Ratio
lvPPA: logopenic variant primary progressive aphasia
MCI: mild cognitive impairment
MGH: Massachusetts General Hospital
nfvPPA: non-fluent/agrammatic variant primary progressive aphasia
PASS SoB: Progressive Aphasia Severity Scale Sum of Boxes
PPA: primary progressive aphasia
PPAOS: primary progressive apraxia of speech
PSP: progressive supranuclear palsy
svPPA: semantic variant Primary Progressive Aphasia
U: Unit
WAB-R: Western Aphasia Battery – Revised

## Acknowledgements

We thank the patients and their families for participating in this research. We thank Inola Howe for assisting in the transcription of the handwritten samples. We thank Dr. Mark Eldaief for contributing with patient neurological examinations. We thank Dr. David Caplan for his recommendations regarding interpretation of the data.

## Funding

S.J.H. performed this work as a behavioral neurology/neuropsychiatry fellow on the Sydney R. Baer Jr. Track of the MGH Behavioral Neurology & Neuropsychiatry Fellowship Program, which is funded by the Sidney R. Baer Jr. Foundation. Additional support for this work was provided by funding from NIH grants R01 DC014296, R21 AG073744, R21 DC019567, and Tommy Rickles Endowed Chair in PPA Research.

## Competing interests

## Notes

### Competing Interest Statement

BCD is a consultant for Acadia, Alector, Arkuda, Biogen, Denali, Eisai, Genentech, Lilly, Merck, Takeda, Wave Lifesciences and receives publishing royalties from Cambridge University Press, Elsevier, Oxford University Press.

### Author Declarations

Ethics committee/IRB of Massachusetts General Hospital gave ethical approval for this work. All participants (and their care partners for patients with PPA) gave written informed consent in accordance with guidelines established by the Mass General Brigham Healthcare System Institutional Review Boards which govern human subjects research at Massachusetts General Hospital.

## References

1. Mesulam MM. Primary progressive aphasia. Annals of neurology. 2001;49(4):425–432.

2. Gorno-Tempini ML, Hillis AE, Weintraub S, et al. Classification of primary progressive aphasia and its variants. Neurology. 2011;76(11):1006–1014.

3. Patterson K, Suzuki T, Wydell T, Sasanuma S. Progressive aphasia and surface alexia in Japanese. Neurocase. 1995;1(2):155–165.

4. Chandregowda A, Duffy JR, Machulda MM, Lowe VJ, Whitwell JL, Josephs KA. Neurodegeneration of the visual word form area in a patient with word form alexia. Neurology and clinical neuroscience. 2021;9(4):359.

5. Utianski RL, Duffy JR, Savica R, Whitwell JL, Machulda MM, Josephs KA. Molecular neuroimaging in primary progressive aphasia with predominant agraphia. Neurocase. 2018;24(2):121–123.

6. Graham NL, Patterson K, Hodges JR. When more yields less: Speaking and writing deficits in nonfluent progressive aphasia. Neurocase. 2004;10(2):141–155.

7. Sitek EJ, Barczak A, Kluj-Kozłowska K, et al. Writing in Richardson variant of progressive supranuclear palsy in comparison to progressive non-fluent aphasia. Neurologia i Neurochirurgia Polska. 2015;49(4):217–222.

8. Code C, Muller N, Tree J, Ball M. Syntactic impairments can emerge later: progressive agrammatic agraphia and syntactic comprehension impairment. Aphasiology. 2006;20(9):1035–1058.

9. Sitek EJ, Barczak A, Kluj-Kozłowska K, Kozłowski M, Barcikowska M, Sławek J. Is descriptive writing useful in the differential diagnosis of logopenic variant of primary progressive aphasia, Alzheimer’s disease and mild cognitive impairment? Neurologia i neurochirurgia polska. 2015;49(4):239–244.

10. Zhou J, Wang J-A, Jiang B, Qiu W-J, Yan B, Wang Y-H. A clinical, neurolinguistic, and radiological study of a Chinese follow-up case with primary progressive aphasia. Neurocase. 2013;19(5):427–433.

11. Heitkamp N, Schumacher R, Croot K, et al. A longitudinal linguistic analysis of written text production in a case of semantic variant primary progressive aphasia. Journal of Neurolinguistics. 2016;39:26–37.

12. Hwang YT, Strikwerda-Brown C, El-Omar H, et al. “More than words”–Longitudinal linguistic changes in the works of a writer diagnosed with semantic dementia. Neurocase. 2021;27(3):243–252.

13. Henry ML, Beeson PM, Alexander GE, Rapcsak SZ. Written language impairments in primary progressive aphasia: a reflection of damage to central semantic and phonological processes. Journal of cognitive neuroscience. 2012;24(2):261–275.

14. Tetzloff KA, Utianski RL, Duffy JR, et al. Quantitative analysis of agrammatism in agrammatic primary progressive aphasia and dominant apraxia of speech. Journal of Speech, Language, and Hearing Research. 2018;61(9):2337–2346.

15. Gallée J, Cordella C, Fedorenko E, et al. Breakdowns in informativeness of naturalistic speech production in primary progressive aphasia. Brain sciences. 2021;11(2):130.

16. Ravid D, Berman RA. Information density in the development of spoken and written narratives in English and Hebrew. Discourse Processes. 2006;41(2):117–149.

17. Dickerson BC, McGinnis SM, Xia C, et al. Approach to atypical Alzheimer’s disease and case studies of the major subtypes. CNS spectrums. 2017;22(6):439–449.

18. Sapolsky D, Domoto-Reilly K, Dickerson BC. Use of the Progressive Aphasia Severity Scale (PASS) in monitoring speech and language status in PPA. Aphasiology. 2014;28(8-9):993–1003.

19. Morris JC. The clinical dementia rating (cdr): Current version and. Young. 1991;41:1588–1592.

20. Nicholas LE, Brookshire RH. A system for quantifying the informativeness and efficiency of the connected speech of adults with aphasia. Journal of Speech, Language, and Hearing Research. 1993;36(2):338–350.

21. Berube S, Nonnemacher J, Demsky C, et al. Stealing cookies in the twenty-first century: Measures of spoken narrative in healthy versus speakers with aphasia. American journal of speech-language pathology. 2019;28(1S):321–329.

22. Rezaii N, Wolff P, Price BH. Natural language processing in psychiatry: the promises and perils of a transformative approach. The British Journal of Psychiatry. 2022;220(5):251–253.

23. Klein D, Manning CD. Accurate unlexicalized parsing. 2003:423–430.

24. Honnibal M, Montani I. spaCy 2: Natural language understanding with Bloom embeddings, convolutional neural networks and incremental parsing. To appear. 2017;7(1):411–420.

25. Graham NL. Dysgraphia in primary progressive aphasia: Characterisation of impairments and therapy options. Aphasiology. 2014;28(8-9):1092–1111.

26. Shim H, Hurley RS, Rogalski E, Mesulam M-M. Anatomic, clinical, and neuropsychological correlates of spelling errors in primary progressive aphasia. Neuropsychologia. 2012;50(8):1929–1935.

27. Neophytou K, Wiley RW, Rapp B, Tsapkini K. The use of spelling for variant classification in primary progressive aphasia: Theoretical and practical implications. Neuropsychologia. 2019;133:107157.

28. Sepelyak K, Crinion J, Molitoris J, et al. Patterns of breakdown in spelling in primary progressive aphasia. Cortex. 2011;47(3):342–352.

29. Tee BL, Kwan-Chen LYL, Chen T-F, et al. Dysgraphia phenotypes in native Chinese speakers with primary progressive aphasia. Neurology. 2022;98(22):e2245–e2257.

30. Graham NL. Dysgraphia in dementia. Neurocase. 2000;6(5):365–376.

31. Lambon Ralph MA, Patterson K. Acquired Disorders of Reading. The Science of Reading: A Handbook. 2005:413–430.

32. Mesulam M-M, Rogalski EJ, Wieneke C, et al. Primary progressive aphasia and the evolving neurology of the language network. Nature Reviews Neurology. 2014;10(10):554–569.

33. Croisile B, Ska B, Brabant M-J, et al. Comparative study of oral and written picture description in patients with Alzheimer’s disease. Brain and language. 1996;53(1):1–19.

34. Wu TQ, Miller ZA, Adhimoolam B, et al. Verbal creativity in semantic variant primary progressive aphasia. Neurocase. 2015;21(1):73–78.

35. Rezaii N, Mahowald K, Ryskin R, Dickerson B, Gibson E. A syntax–lexicon trade-off in language production. Proceedings of the National Academy of Sciences. 2022;119(25):e2120203119.

36. Clarke N, Foltz P, Garrard P. How to do things with (thousands of) words: Computational approaches to discourse analysis in Alzheimer’s disease. Cortex. 2020;

37. Themistocleous C, Neophytou K, Rapp B, Tsapkini K. A tool for automatic scoring of spelling performance. Journal of Speech, Language, and Hearing Research. 2020;63(12):4179–4192.

38. Mazzeo S, Polito C, Lassi M, et al. Loss of speech and functional impairment in Alzheimer’s disease-related primary progressive aphasia: predictive factors of decline. Neurobiology of Aging. 2022;

39. Semler E, Anderl-Straub S, Uttner I, et al. A language-based sum score for the course and therapeutic intervention in primary progressive aphasia. Alzheimer’s research & therapy. 2018;10(1):1–10.

